# Estimation of local novel coronavirus (COVID-19) cases in Wuhan, China from off-site reported cases and population flow data from different sources

**DOI:** 10.1101/2020.03.02.20030080

**Authors:** Zian Zhuang, Peihua Cao, Shi Zhao, Yijun Lou, Shu Yang, Weiming Wang, Lin Yang, Daihai He

## Abstract

**Backgrounds:** In December 2019, a novel coronavirus (COVID-19) pneumonia hit Wuhan, Hubei Province, China and spread to the rest of China and overseas. The emergence of this virus coincided with the Spring Festival Travel Rush in China. It is possible to estimate total number of cases of COVID-19 in Wuhan, by 23 January 2020, given the cases reported in other cities and population flow data between cities.

**Methods:** We built a model to estimate the total number of cases in Wuhan by 23 January 2020, based on the number of cases detected outside Wuhan city in China, with the assumption that if the same screening effort used in other cities applied in Wuhan. We employed population flow data from different sources between Wuhan and other cities/regions by 23 January 2020. The number of total cases was determined by the maximum log likelihood estimation.

**Findings:** From overall cities/regions data, we predicted 1326 (95% CI: 1177, 1484), 1151 (95% CI: 1018, 1292) and 5277 (95% CI: 4732, 5859) as total cases in Wuhan by 23 January 2020, based on different source of data from Changjiang Daily newspaper, Tencent, and Baidu. From separate cities/regions data, we estimated 1059 (95% CI: 918, 1209), 5214 (95% CI: 4659, 5808) as total cases in Wuhan in Wuhan by 23 January 2020, based on different sources of population flow data from Tencent and Baidu.

**Conclusion:** Sources of population follow data and methods impact the estimates of local cases in Wuhan before city lock down.

## Introduction

In December 2019, a cluster of patients with pneumonia of unknown causes were reported in Wuhan, Hubei Province, China [1]. On 7 January 2020, a novel coronavirus, named COVID-19, was identified as the cause of this outbreak [2]. This COVID-19 virus shares the common characteristics of coronavirus and is expected to become more virulent when establishing efficient human-to-human transmission [3]. The emergence of this virus coincided with the Spring Festival Travel Rush in China. It was estimated that there would be around 3 billion trips made in China during the period of 10 January to 18 February 2020 [4]. Some researchers have pointed out the risk of regional and global disease spreading during the Spring Festival Travel Rush [5]. However, due to the small number of severe patients reported by mid-January and most cases were linked to the Huanan Seafood Market of Wuhan city, neither international nor regional travelling restrictions were implemented to Wuhan at the early stage of this outbreak. On 13 January 2020, the first exported case from Wuhan was reported in Thailand and the case numbers dramatically increased after the diagnostic kits became available in mid-January. As of 23 February 2020, there were 77042 laboratory confirmed cases and 2445 deaths (46.93% and 75.91% in Wuhan) [6]. In recognition of a wide-spreading outbreak, the government has suspended all public transportations inside Wuhan city since 23 January 2020, and some regional travelling restrictions were also implemented by other cities [3].

### Objective

At early stage of this outbreak, the cases might have been seriously underreported due to the lack of diagnostic kits and insufficient screening for all suspected cases. Several efforts have been made to estimate the case numbers using different modelling approaches, and the estimates range from 1732 to 4000 during the period of 17-20 January [7,8,9].

In this study, we aimed to estimate the number of COVID-19 cases in Wuhan, utilizing the cases exported to other large cities of mainland China and different sources of the population flow data between Wuhan and these cities. The estimates were made by 23 January 2020 (before the suspension of public transportations in Wuhan). We assumed that the exported cases were less likely underreported, as stringent temperature screening was implemented at airports and railway stations. We compare these estimates to daily numbers of confirmed cases exported from Wuhan in surveillance data to evaluate the extent of underreporting.

### Data

We obtained daily number of inbound and outbound domestic passengers travelling by air, train or road to/from Wuhan from three data sources:

1. Tencent’s LBS (location-based services) database (see: https://heat.qq.com/). According to location data of Tencent’s mobile software users, population flow number during 10 December 2016 and 24 January 2017 was generated, between Wuhan and twenty-five cities/regions (Anhui, Beijing, Chongqing, Fujian, Gansu, Guangdong, Guangxi, Guizhou, Hainan, Hebei, Heilongjiang, Henan, Hunan, Hubei(Outside Wuhan), Jiangsu, Jiangxi, Jilin, Liaoning, Ningxia, Shandong, Shanghai, Sichuan, Tianjin, Yunnan, Zhejiang). Please note that the cities in Hubei here means all cities in Hubei other than Wuhan.
2. Baidu map database (see: https://qianxi.baidu.com/). According to location data of Baidu’s mobile software users, population flow number from 1 to 20 January 2020 was generated, between Wuhan and twenty-seven cities/regions (Anhui, Beijing, Chongqing, Fujian, Gansu, Guangdong, Guangxi, Guizhou, Hainan, Hebei, Heilongjiang, Henan, Hunan, Hubei(Outside Wuhan), Jiangsu, Jiangxi, Jilin, Liaoning, Ningxia, Shandong, Shanghai, Shanxi, Shannxi, Sichuan, Tianjin, Yunnan, Zhejiang).
3. The news platform. It has reported that there were 4.1 million outbound passengers from Wuhan in the first ten days of the Spring Festival Travel Rush (10-19 January 2020) via railways, highways and airways [10].

We equally divided the data to get an average daily population flow.

As shown in Fig 1, we also collected daily numbers of exported cases from Wuhan to other cities in China, and all secondary cases of family or hospital clusters were excluded from analysis [11]. Eight cases from Guangdong were excluded due to the lack of traveling history to Hubei prior to illness onset. As for rest 371 cases that not specified as secondary case, we assume that the probability of a single case being an imported case is *θ*, and each case is independent from each other. Then all of these unspecified cases follow a binomial distribution (n, *θ*), where *θ* represents the probability that a case is exported from Wuhan. Since the most cases detected outside Wuhan are imported cases, by 23 January 2020 [12], we estimated daily numbers of imported COVID-19 cases from Wuhan based on different level of probability *θ* (1,0.9,0.8), see Table S1 in appendix.

**Figure 1:**
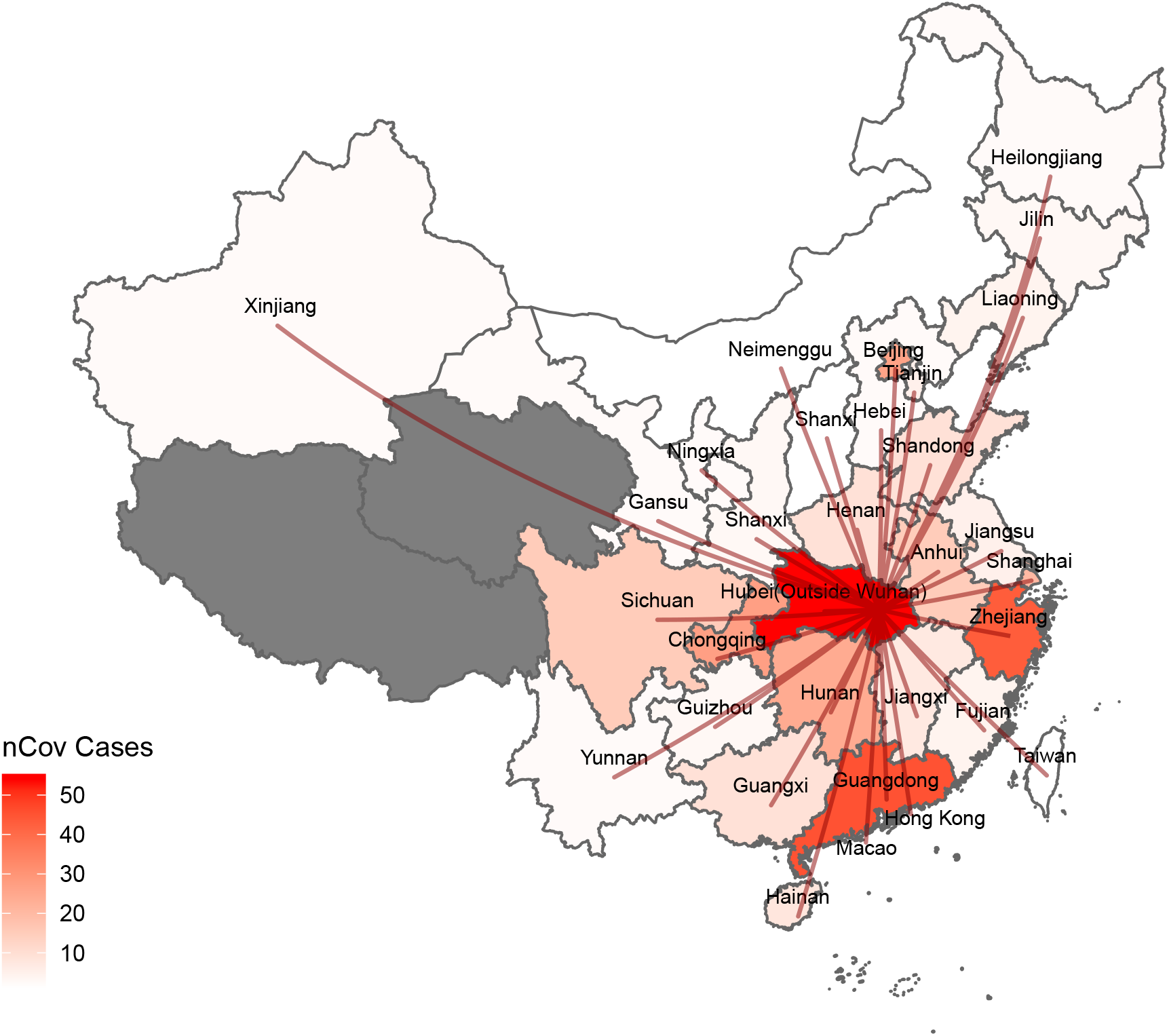
The geographical distribution of imported COVID-19 in China. This figure reported COVID-19 cases outside Wuhan (excluded 8 from Guangdong), the dark grey area indicates the regions with zero COVID-19 cases as of 23 January 2020. Red paths show routes from Wuhan to other cities/regions.

## Methods

Daily number of diagnosed cases of COVID-19 infection in China was assumed to follow a Binomial distribution [7], as in Eqn (1), where *n* is total number of cases and *p* is probability of finding any cases overseas.

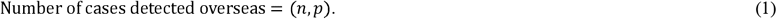

The probability *p* can be derived from dividing daily outbound international passengers of Wuhan by the population size that the Wuhan airport serves and multiplying by the average time of case discovery, see Eqn (2).

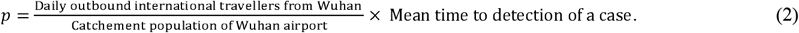

Similarly, we used exported cases from Wuhan to predict numbers of local imported COVID-19 cases. We assumed a population of 19 million (catchment population) travelling through the airport, railway stations and highways in Wuhan, and a 10-day delay on average, which accounted for the time interval between the timings of infection and case reported [7]. Furthermore, it was assumed that all cases in other cities/regions outside Wuhan are detected. If cases are missed in other cities/regions, our forecast will underestimate the real number of cases.

Apart from predict number of COVID-19 cases through overall data, we also tried to forecast number of COVID-19 cases base on separate city/region’s data since more details may convey more information. A parameter λ was set as total COVID-19 cases. Based on the data obtained from each city/region, we estimated the λ, which is the most likely to make these results appear by calculating maximum of log-likelihood.

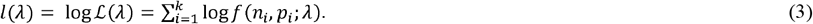

In Equation (3), *l*(·) is the total log-likelihood. The k represents the total number of cities/regions calculated. The *n, p* represents the number of cases reported overseas and probability of finding any cases overseas respectively. After obtaining λ, since residuals of log maximum likelihood estimation follows Chi-square distribution [13], 95% confidence intervals (95% CI) of log-likelihood, *l*, can be calculated. Then we can extrapolate a 95% CI about COVID-19 cases.

In addition, we analyze the correlation between the two data sources. We found that Pearson Correlation Coefficient of Baidu and Tencent data is 0.9943 (95% CI: 0.9871, 0.9976). We assumed a linear relation between the Baidu data and Tencent data, see Eqn (4). Then we tested null hypothesis *H*_0_ that *α* = 0.

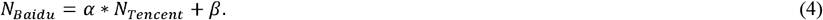

## Result

As for separate cities/regions data, based on different data sources, we summed the total population flow from Wuhan to other cities/regions and the total number of cases in those cities. Then we estimate total number and 95% confidence interval (95% CI) about COVID-19 cases. We predicted 1326 (95% CI: 1177, 1484), 1151 (95% CI: 1018, 1292) and 5277 (95% CI: 4732, 5859) as total cases in Wuhan by 23 January, based on different source of data, see Table 1.

**Table 1:** Summary table of the outbreak size estimates in Wuhan, China, 23 January 2020, through overall data.

|  | News Data |  |  | Tencent Data |  |  | Baidu Data |  |  |
| --- | --- | --- | --- | --- | --- | --- | --- | --- | --- |
| | Baseline<br>($\theta=1$) | ($\theta=0.9$) | ($\theta=0.8$) | Baseline<br>($\theta=1$) | ($\theta=0.9$) | ($\theta=0.8$) | Baseline<br>($\theta=1$) | ($\theta=0.9$) | ($\theta=0.8$) |
| **Estimated total number of cases (95% CI)** | 1691<br>(1542, 1849) | 1548<br>(1405, 1699) | 1353<br>(1220, 1495) | 1507<br>(1374, 1648) | 1363<br>(1237, 1497) | 1202<br>(1084, 1329) | 5637<br>(5092, 6219) | 5105<br>(4587, 5660) | 4494<br>(4009, 5016) |
| **Estimated number of cases in Wuhan (95% CI)** | 1326<br>(1177, 1484) | 1214<br>(1071, 1365) | 1061<br>(928, 1203) | 1151<br>(1018, 1292) | 1043<br>(897, 1157) | 917<br>(799, 1044) | 5277<br>(4732, 5859) | 4781<br>(4263, 5336) | 4206<br>(3721, 4728) |

As for separate cities/regions data, we estimated total number of cases, λ, by using the maximum log-likelihood estimation (MLE) approach based on the population flow data from Tencent and Baidu, see Fig 2 (a,b). Then we estimated 95% confidence interval (95% CI) about COVID-19 cases. We predicted 1059 (95% CI: 918, 1209), 5214 (95% CI: 4659, 5808) as total cases in Wuhan by 23 January 2020, based on different sources of population flow data, see Table 2.

**Table 2:** Estimating the outbreak size in Wuhan, China, 23 January 2020, through separate cities/regions data

|  | Tencent Data |  |  | Baidu Data |  |  |
| --- | --- | --- | --- | --- | --- | --- |
| | Baseline<br>($\theta=1$) | ($\theta=0.9$) | ($\theta=0.8$) | Baseline<br>($\theta=1$) | ($\theta=0.9$) | ($\theta=0.8$) |
| **Estimated total number of cases** | 1415<br>(1274, 1565) | 1279<br>(1146, 1422) | 1128<br>(1003, 1263) | 5574<br>(5019, 6168) | 5047<br>(4520, 5613) | 4444<br>(3950, 4976) |
| **Estimated number of cases in Wuhan (95% CI)** | 1059 (918, 1209) | 959 (826, 1120) | 843 (718, 978) | 5214<br>(4659, 5808) | 4723<br>(4196, 5289) | 4156<br>(3662, 4688) |

**Figure 2:**
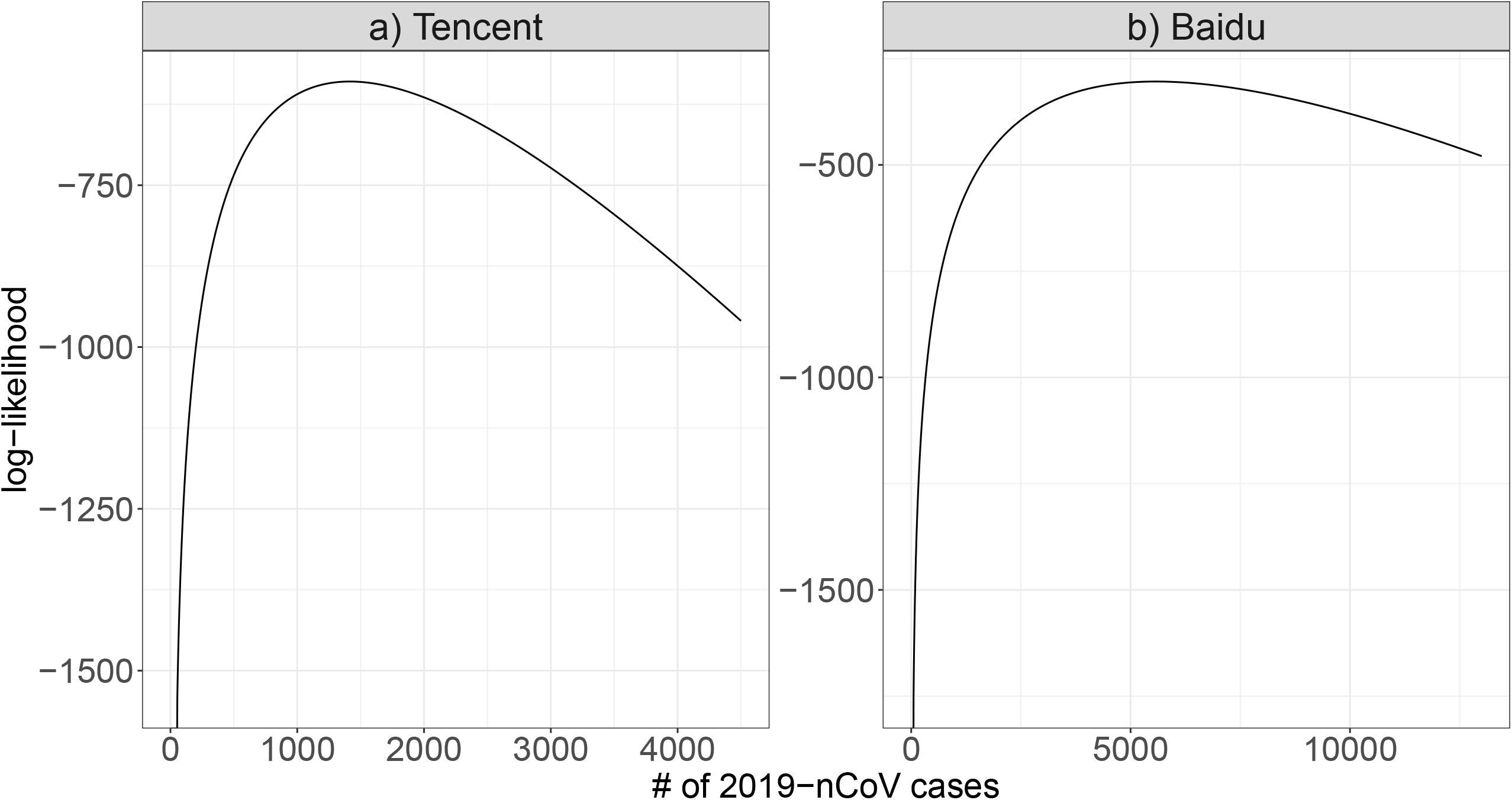
Log maximum likelihood estimation for λ.

In addition, as for the correlation analysis between data from different sources, we got result that estimated coefficient *α* equals 0.23, *β* equals 689.11 and *p-value* was less than 2e-16. Then we rejected the null hypothesis *H*_0_ under 99.99% CI, which suggests that two sources of data are likely to have a linear relation.

## Discussion

Recent study by Imai *et al*. estimated that a total of 4000 (95% CI: 1000-9700) cases in 18 January 2020 [8] and Leung *et al*. estimated that 1343 (95% CI: 547-3446) cases had onset of symptoms in Wuhan during 1-17 January 2020 [9]. In general, compared with the total number of confirmed cases provided by the government, which is 495 [14], Imai *et al*. and Leung *et al*. predicted larger number of cases [7,9]. This is partly because the screening effort targeting population from Wuhan in other cities is much more effective than the local screening effort in Wuhan due to the worsen situation. Based on three different sources of population flow data, we generated the total COVID-19 cases prediction closest to the official report through Tencent data. Baidu Data provided closest result to the forecast by Imai et al. [8]. Meanwhile, in sensitivity analysis, Table 1 and Table 2 show that slight fluctuations of probability that a case is exported from Wuhan will have little impact on the forecast.

Estimates of the population outflow provided by news, Baidu and Tencent show substantial fluctuation, resulting in wide predictions. We found that Baidu and Tencent data show significant linear relation, which verified each other that the general pattern of data is reasonable. After simple linear transformation, two sets of data become similar (see Fig 3). One possible reason for the phenomenon is that different institutions have various definition about the number of people flow form one city to another. Methods includes people who travel to other city through Wuhan in the population flow may provide much larger figure than those only calculate people only originally depart from Wuhan. At the same time, multiple round trips may also affect the count. Another possible reason is that Baidu and Tencent would fail to track whole amount of population flow since not everyone uses mobile phone software from Baidu and Tencent.

**Figure 3:**
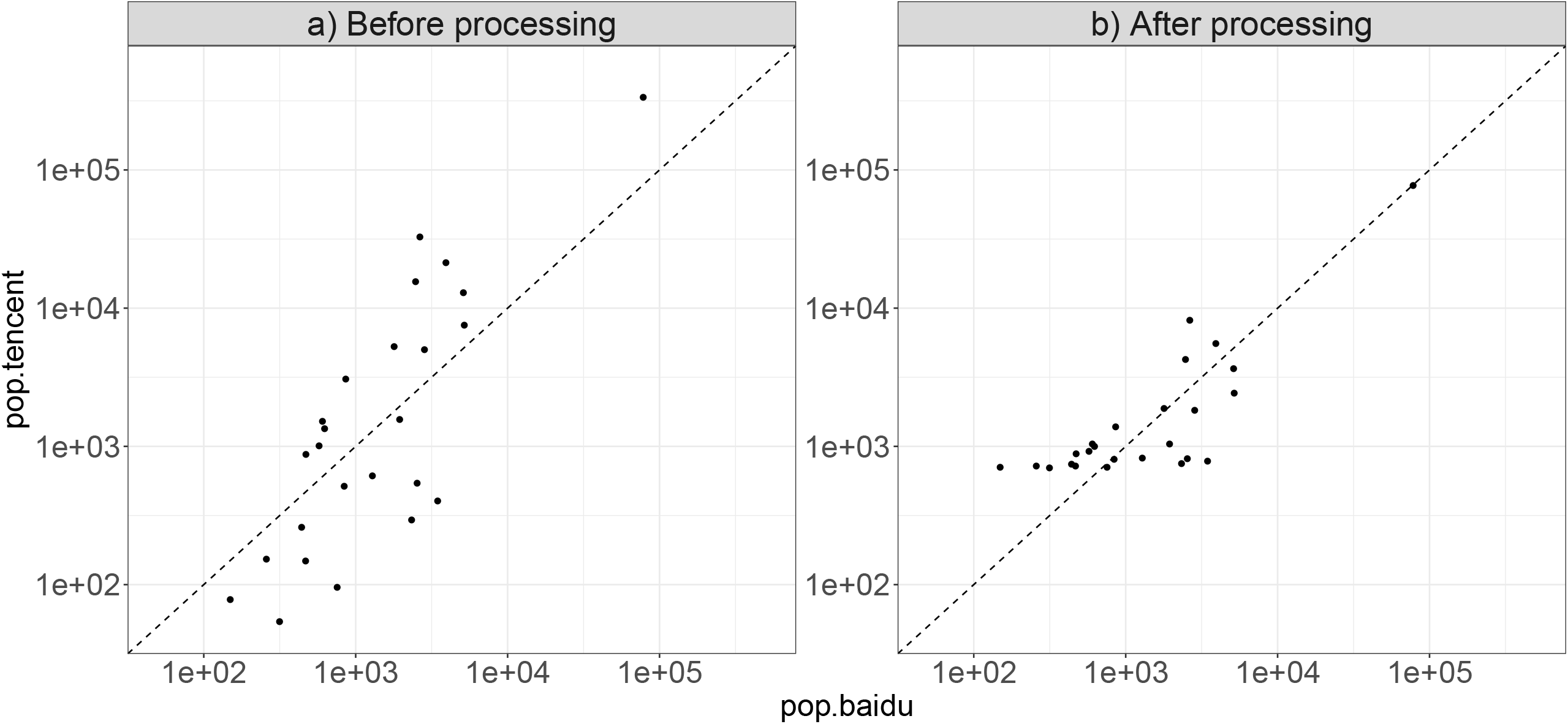
Processing data from Baidu and Tencent. X-axis presents the number of population flow between Wuhan and other cities/regions from Tencent data. Y-axis presents the number of population flow between Wuhan and other cities/regions from Baidu date.

Imai et al. suggested that by further improving the definition and testing of COVID-19 cases, and further expanding the scope of epidemic monitoring, the gap between the estimated number and official reported cases will be further narrowed. According to our results, statistics of population flow also play significant roles in prediction. At present, many researches use data from Baidu and Tencent platforms [9,15,16]. It is not clear which data source can be used to make the most accurate prediction yet, while it is possible for forecasts based on a single data source to be inaccurate. To sum up, it is necessary to make a comprehensive analysis based on different statistics before reaching any conclusions.

## Conclusion

Population flow data were used in estimating the size of the epidemic of COVID-19 in Wuhan, China before the lock down. We found that population flow data from different sources may lead to significantly different estimates, although all estimates suggested much larger sizes than officially reported. Using cases reported off-site to estimate the size (course) of the epidemic in the epicenter is a common technique and will be used in future epidemics, especially when the epicenter lacks medical resources thus cases are under-reported. We argue that reliability of estimates of population flow data should be taken into consideration.

## Data Availability

We used publicly available data only.

## Declarations

### Ethics approval and consent to participate

The ethical approval or individual consent was not applicable.

### Availability of data and materials

All data and materials used in this work were publicly available.

### Consent for publication

Not applicable.

## Acknowledgements

None.

## Disclaimer

The funding agencies had no role in the design and conduct of the study; collection, management, analysis, and interpretation of the data; preparation, review, or approval of the manuscript; or decision to submit the manuscript for publication.

## Authors’ Contributions

All authors conceived the study, carried out the analysis, discussed the results, drafted the first manuscript, critically read and revised the manuscript, and gave final approval for publication.

## Appendix

**Table S1:** COVID-19 cases used in this paper (for different values of p).

| City | p=1 | p=0.9 | p=0.8 |
| --- | --- | --- | --- |
| Zhejiang | 43 | 39 | 34 |
| Shanghai | 20 | 18 | 16 |
| Guangdong | 45 | 41 | 36 |
| Shanxi | 3 | 3 | 2 |
| Hainan | 8 | 7 | 6 |
| Heilongjiang | 4 | 4 | 3 |
| Jiangxi | 7 | 6 | 6 |
| Guizhou | 3 | 3 | 2 |
| Anhui | 15 | 14 | 12 |
| Chongqing | 27 | 24 | 22 |
| Hunan | 24 | 22 | 19 |
| Liaoning | 4 | 4 | 3 |
| Fujian | 5 | 5 | 4 |
| Jilin | 3 | 3 | 2 |
| Sichuan | 15 | 14 | 12 |
| Shandong | 9 | 8 | 7 |
| Guangxi | 13 | 12 | 10 |
| Beijing | 26 | 23 | 21 |
| Ningxia | 2 | 2 | 2 |
| Gansu | 2 | 2 | 2 |
| Jiangsu | 9 | 8 | 7 |
| Henan | 9 | 8 | 7 |
| Shanxi | 1 | 1 | 1 |
| Tianjin | 5 | 5 | 4 |
| Yunnan | 2 | 2 | 2 |
| Hubei (exclude wuhan) | 54 | 49 | 43 |
| Hebei | 2 | 2 | 2 |

*Table S1: COVID-19 cases used in this paper (for different values of $p$ ).*
|  |  |  |  |
| --- | --- | --- | --- |
| Taiwan | 1 | 1 | 1 |
| Hong Kong | 2 | 2 | 2 |
| Macao | 2 | 2 | 2 |
| **Total** | **365** | **334** | **292** |

